# Modelling the impacts of public health interventions and weather on SARS-CoV-2 Omicron outbreak in Hong Kong

**DOI:** 10.1101/2022.05.25.22275487

**Authors:** Hsiang-Yu Yuan, Jingbo Liang

## Abstract

**Background:** Hong Kong, has operated under a zero-Covid policy in the past few years. As a result, population immunity from natural infections has been low. The ‘fifth wave’ in Hong Kong, caused by the Omicron variant, grew substantially in February 2022 during the transition from winter into spring. The daily number of reported cases began to decline quickly in a few days after social distancing regulations were tightened and rapid antigen test (RAT) kits were largely distributed. How the non-pharmaceutical interventions (NPIs) and seasonal factors (temperature and relative humidity) could affect the spread of Omicron remains unknown.

**Methods:** We developed a model with stratified immunity, to incorporate antibody responses, together with changes in mobility and seasonal factors. After taking into account the detection rates of PCR test and RAT, we fitted the model to the daily number of reported cases between 1 February and 31 March, and quantified the associated effects of individual NPIs and seasonal factors on infection dynamics.

**Findings:** Although NPIs and vaccine boosters were critical in reducing the number of infections, temperature was associated with a larger change in transmissibility. Cold days appeared to drive *Re* from about 2–3 sharply to 10.6 (95%CI: 9.9–11.4). But this number reduced quickly below one a week later when the temperature got warmer. The model projected that if weather in March maintained as February’s average level, the estimated cumulative incidence could increase double to about 80% of total population.

**Interpretation:** Temperature should be taken into account when making public health decisions (e.g. a more relaxed (or tightened) social distancing during a warmer (or colder) season).

## Introduction

China, Hong Kong, Taiwan, New Zealand have been or previously been operating under a zero-Covid policy, likely resulting in a highly susceptible population due to low levels of previous virus exposure. The recent Omicron variant was estimated to cause a high infection rate with substantial hospitalisations and deaths in such countries, if the outbreak was not controlled [1]. Successfully managing the outbreaks in such highly susceptible populations is a challenging task.

The SARS-CoV-2 Omicron outbreak (known as the ‘fifth wave’) in Hong Kong, originally linked to imported cases [2], was exacerbated since the Chinese Spring Festival, starting on 1 February 2022. This wave resulted in more than 1.1 million confirmed cases (in a total population of 7.48 million) within two months. Despite the social distancing was soon tightened on 10 February 2022 [3], the outbreak continued to grow. The number of new cases suddenly rose from about 10,000 on 25 February 2022 to the peak of near 80,000 less than a week. Meanwhile on 25 February, because the PCR testing and contact-tracing systems were overwhelmed, the Government decided that Rapid Antigen Test (RAT) can be used for case confirmation and began to distribute the kits [4][5]. Surprisingly, several days later, the number reduced rapidly. The sharp pattern of the epidemic was clearly different from the slow plateau-like infections that were commonly seen in many nearby countries around the same time (Figure S1). Modelling how this outbreak was managed in such an agile way allows us to formulate recommendations for the management of the future outbreaks.

Faced with a growing number of cases, vaccine boosters received increased from approximately 20,000 to over 40,000 per day since February 2022 [6]. The distribution of neutralizing antibody titres induced by vaccine against the Omicron variant was observed in Hong Kong, which was not particularly high [7]. A higher titre level can reduce the susceptibility of infection more [8]. To take into account differences in antibody responses, epidemic models with multiple susceptible states (called stratified immunity) have been developed [9], in which each titre level was mapped to one of the states. Modelling stratified immunity allows a more natural description of the population immunity. The timing and the magnitude of the epidemic dynamics can be reconstructed more accurately [9].

Besides NPI and vaccination, the spread of COVID-19 is postulated to be influenced by temperature and humidity. Lower temperatures have been found to be associated with higher transmissibility [10][11][12]. In addition, the relationship between relative humidity and the transmissibility has also been studied [12][13][14]. A recent study suggested incorporating temperature can improve the accuracy of model forecasts [10]. As Hong Kong experienced cooler days than usual with high relative humidity during February 2022 and the spring season brought warmer conditions in the following months [15], these factors should be modelled.

In order to understand the drivers of the fifth wave and how this was successfully managed, a model embedding with stratified immunity [9] and non-pharmaceutical interventions (NPIs) [16] was developed. We aimed at quantifying the associated impacts of individual NPIs, vaccine booster and other seasonal factors (i.e. temperature and humidity) after incorporating daily changes of relevant data.

## Material and Methods

Daily vaccination rates of BioNTech and CoronaVac were collected from the COVID-19 Thematic Website [3]. Mobility data were collected from Google mobility [17]. Daily mean temperature and relative humidity were collected from Hong Kong Observatory [18]. Daily number of reported cases detected by either PCR or RAT were collected from the Hong Kong Centre for Health Protection [19].

The period of the fifth wave in our study was defined as beginning when infected cases are consistently above 100 cases on 1 February 2022 and as ending on 31 March 2022 when the daily case number has been constantly less than 10% of the epidemic peak since then. In order to capture the impacts of NPIs and seasonal factors in a population with changing immunity, we extended our previous deterministic stratified immunity model [9] to incorporate daily changes in vaccination, mobility and weather conditions (Figure 1 and Figure S2).

**Figure 1.**
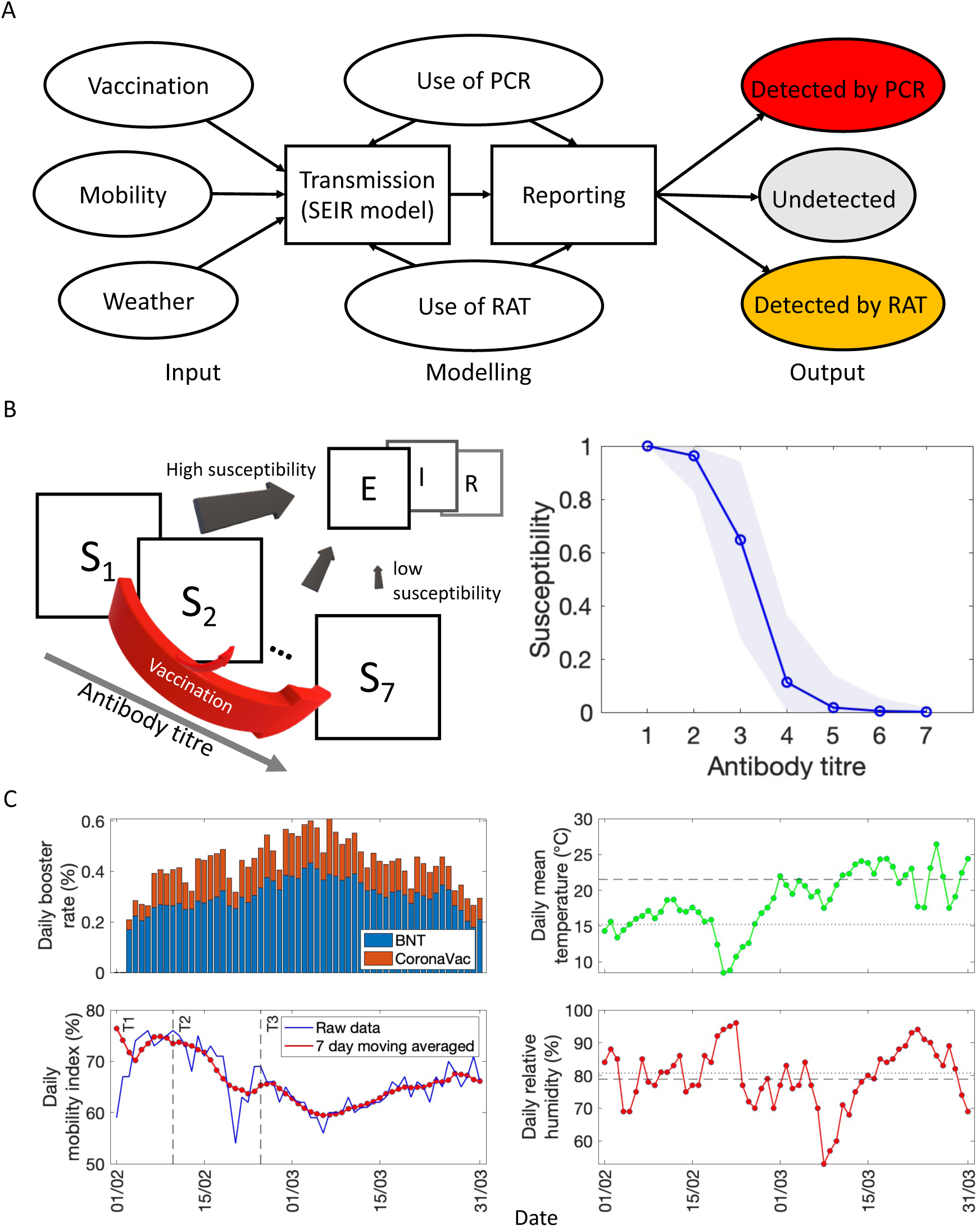
Daily uncertainties of social distancing, vaccination and weather conditions were considered in the modelling process. (A) Factors that affect the number of reported cases detected, including daily uncertainties (i.e. vaccination, mobility and weather) and the uses of PCR and RAT. Reporting rates and delays are taken into account to allow the model output to compare to actual reported cases, detected by PCR (red) and RAT (yellow). (B) Left, modelling the change of susceptible individuals with different antibody titre levels after vaccination. *S*_*i*_ represents the susceptible individuals having *i*th antibody titre level. Right, the relationship between antibody titre and susceptibility. (C) Daily booster vaccine rates, mobility, and weather conditions (such as daily mean temperature and relative humidity) throughout the outbreak. *Left*, T1-3 represent individual social distancing tightenings. *Right*, dotted lines represent average weather conditions in February and dashed lines represents average values in March.

The force of infection *λ*_*i*_ of individuals having *i*th antibody titre level is proportional to their susceptibility, social mixing, and temperature and relative humidity they were exposed to at each day:

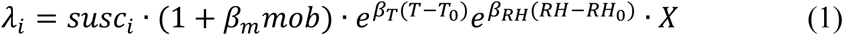

where *susc*_*i*_ is the susceptibility of infection for susceptible individuals having antibody titre level *i. β*_*m*_ is the coefficient for the percent reduction of population mobility (*mob*), compared to the pre-pandemic period. *T* is the daily temperature and *T*_0_ is the baseline temperature (i.e. the average temperature in February). *β*_*T*_ is the coefficient for temperature. Similarly, *β*_*RH*_ is the coefficient for relative humidity. *RH* is the daily relative humidity and *RH*_0_ is the baseline relative humidity. *X* here represents the effects from other factors, including other NPIs and the number of infected cases (see Supplementary Materials for detailed specification).

### Vaccine-induced protection

The increase of antibody titre due to two different vaccines (BioNTech and CoronaVac are two available vaccines in Hong Kong) resulting from full immunization (2nd dose) or vaccine booster doses was collated from serum data in a previous study [7]. There are seven titre levels (from 1 to 7), indicating different dilutions, such as <1:10, 1:10, 1:20,…, 1:160, and ≥1:320, measured as the highest serum dilution that resulted in >50% reduction in the number of virus plaques (PRNT50) [20]. Antibody boosting was represented as the increase in antibody titre from the original to a boosted level, parameterised by a log-normal distribution (see Figure S3).

The susceptibility of individuals at a particular antibody titre can be described using a sigmoid function (see Figure 1B):

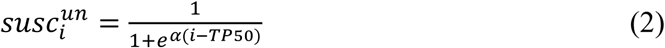

where 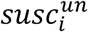 is the probability (un-normalized) of developing disease for individual with antibody titer at level *i* given a contact; *TP*50 is the titre level found to exhibit 50% protection, which was found to be 1:25.6 in a previous study [7]; and α is the shape parameter, which can be estimated by the model. Note that this ‘un-normalized’ probability does not lead to 100% susceptibility when the titre level is at the minimum level. This potentially leads to multi-collinearity issue during model fitting. To avoid this, we further normalized the susceptibility to allow the lowest titre to be fixed at 100%. For the description of the full model, see Supplementary Materials.

We assumed that antibody levels in people who received vaccination more than three months ago had waned already and the amount of antibodies increased 7 days after having the second or third dose [21–23]. Hence, daily booster rate was defined as the proportion of total population who were having the booster dose 7 days ago. Pre-existing immunity was defined as the proportion of individuals who had been vaccinated either with two or three doses between 25th October 2021 and 25th January 2022 (a week before the beginning of the fifth wave) (see Figure 1A and Figure 3A).

### Modelling testing, tracing and isolation

We modelled both types of tests: the PCR test and the RAT. A certain amount of infected cases performed self-testing using RAT. After the latent period, once they were detected, they self-reported positive outcomes (e.g. through an online self-reporting website in Hong Kong), and self-isolated at home. Home-isolated cases were still able to transmit the virus but with a lower rate of 10.9% (95%CI: 7.1– 14.7%) [16] than infectious cases who were not quarantined or isolated. We assumed that the proportion of infected cases that are detected (i.e. detection rate) by PCR was inversely associated with the ‘true’ number of cases (i.e. both detected and undetected cases) following an exponential curve. The average delay between symptom onset and hospitalization was set to 6 days after checking previous reports. Additional delays in reporting PCR-confirmed cases were dependent on the true case number. The delay became longer but changed less when the number of cases was larger. We modelled contact tracing following our previous approach [16]. According to the report from Centre of Health Protection, the average proportion of cases that were identified through contact tracing was reduced from about 50% in January to 10% in February (Figure S4). Therefore, the proportion of contact-traced was set to be 10% in this study. These contact-traced cases were assumed to be either quarantined or isolated at home. For the description of the full model, see Supplementary Materials.

### Model fitting

The posterior distributions of the parameters of the model for Hong Kong were obtained after fitting the model to the daily numbers of reported cases detected by PCR or RAT. The posterior distributions were estimated using a Markov chain Monte Carlo (MCMC) algorithm with 10^6^ steps to guarantee an effective sample size (ESS) of greater than 1000 for all parameters (see Supplementary Materials).

### Calculating effective reproduction number

The time-dependent effective reproduction number, *Re*, was calculated using the next-generation matrix approach after the posterior distributions of the model parameters were obtained [6].

## Results

In order to quantify how the factors driving and controlling the fifth wave in Hong Kong, we modelled the separate effects of NPIs, vaccination and weather in a highly susceptible population.

### Characterising the fifth wave

We first compared two models: the full model, in which the force of infection for susceptible individuals was determined by vaccine-induced protectiveness, the implementation of social distancing, and weather conditions (temperature and relative humidity); and the ‘reduced’ model, in which the weather effects were not included. After estimating the detection rates of PCR and RAT (Figure 1A and Figure S2), only the full model successfully reproduced the pattern of the actual reported case number (Figure 2A,C), i.e. a rapidly increasing trend, followed by a sudden reduction. The reduced model produced a longer plateau. The model performance, measured by deviance information criterion (DIC), was significantly improved when weather conditions were included (1995.1 versus 2650.9). Hence, the full model was used to characterise the fifth wave.

**Figure 2.**
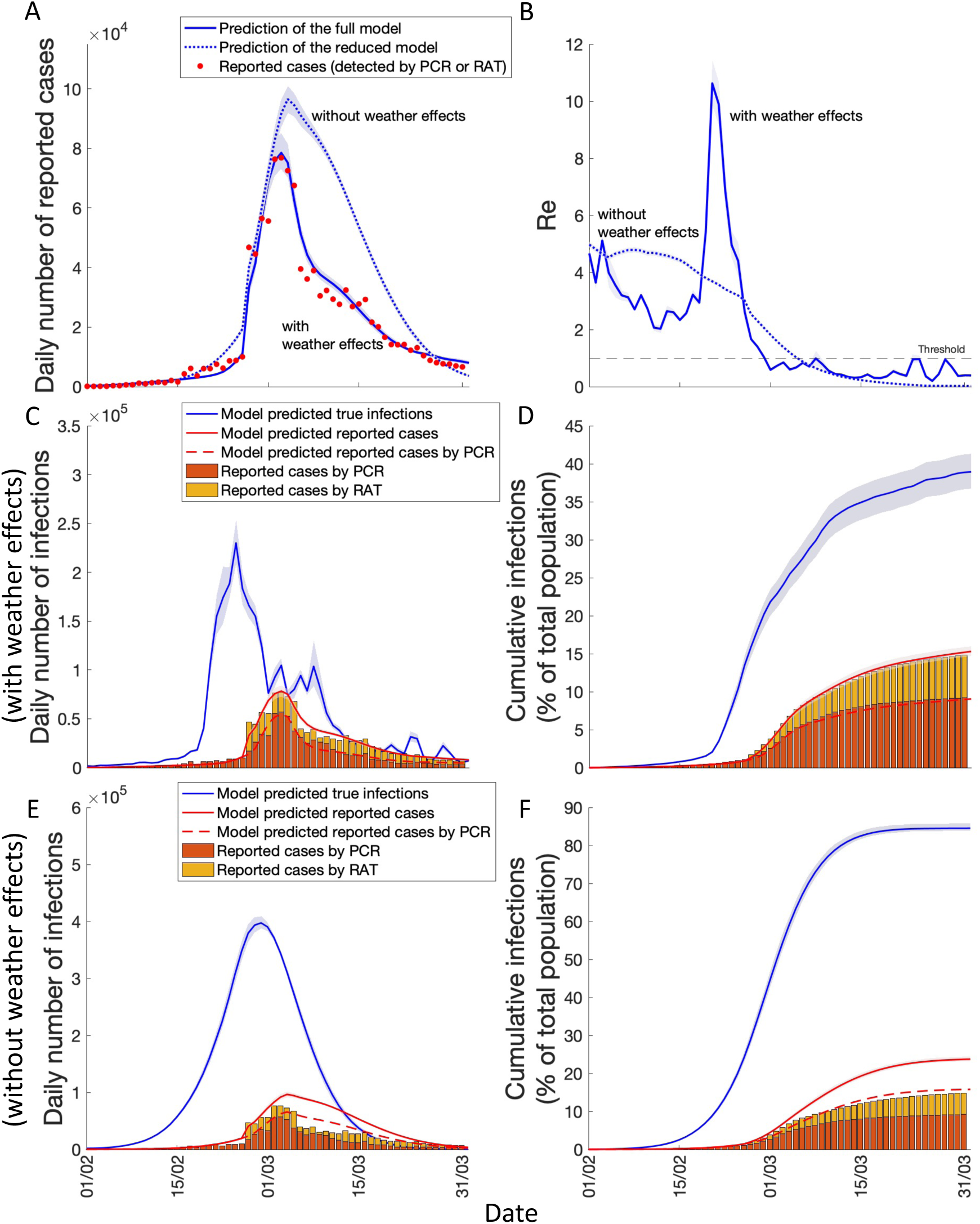
Predicted COVID-19 transmission dynamics of the fifth wave. (A) Predicted numbers of reported cases by the full model (solid blue line) and the reduced model (dotted blue line). (B) Corresponding *Re* of the two models over time. (C) Daily number of true infections (including undetected cases) predicted by the full model. Solid blue line represents the predicted number of daily infections. Red solid line represents the predicted number of daily reported cases. Red dashed line represents the predicted number of daily reported cases detected by PCR. Bars represent the daily numbers of reported cases detected by PCR (red) and RAT (yellow). (D) Cumulative infections (defined as the number of cumulative infections divided by total population size). Individual lines and bars represent the data as (C) but as a percentage of the total population. (EF), same as (C) and (D) but the numbers are predicted by the reduced model.

The maximum daily number of reported cases (76,937) was successfully predicted on 3 March. Furthermore, the model predicted that the true daily number of infections (including the undetected cases) at the time of virus exposure reached its peak on 23 February with the highest estimated of 231,381 (95%CI: 210,705–256,860) and a cumulative 10.5% of the population have been infected (Figure 2CD). The amount of naturally infected cases appeared to be lower than the expected population immunity to suppress this outbreak [24]. However, few days later, the predicted number showed rapid decreases from the peak two times to approximately 80–100 thousand between 27 February – 7 March and to below 40 thousand after 10 March (Figure 2C). The model estimated that up to the end of March, the cumulative incidence was 39.0% (95%Credible Interval (CI): 36.7–41.1%) of population (Figure 2B), while only 38% of them were reported. Nearly 35% of reported cases were detected by RAT (Figure 2D).

*Re* gradually decreased from about 5 to 2-3 in the first three weeks of February but increased sharply to 10.6 (95%CI: 9.9–11.4) on 20 February and maintained few days. The number then reduced quickly to be lower than 1 after just a week in the end of February (Figure 2B).

### Changes in vaccine-induced population immunity

We first assessed whether vaccination was able to explain the rapid reduction in transmissibility during the late February. After incorporating antibody responses of second and third doses of BioNTech and CoronoVac, the pre-existing immunity by 1 February only produced very limited protection (Figure 3A). Our model estimated that the susceptibility (i.e. the probability of being infected given a contact) of individuals following a sigmoid curve as the amount of antibody grew (see Figure 1B). About 99% of individuals whose antibody titre levels were correlated with susceptibility greater than 50% (i.e. <1:40).

**Figure 3.**
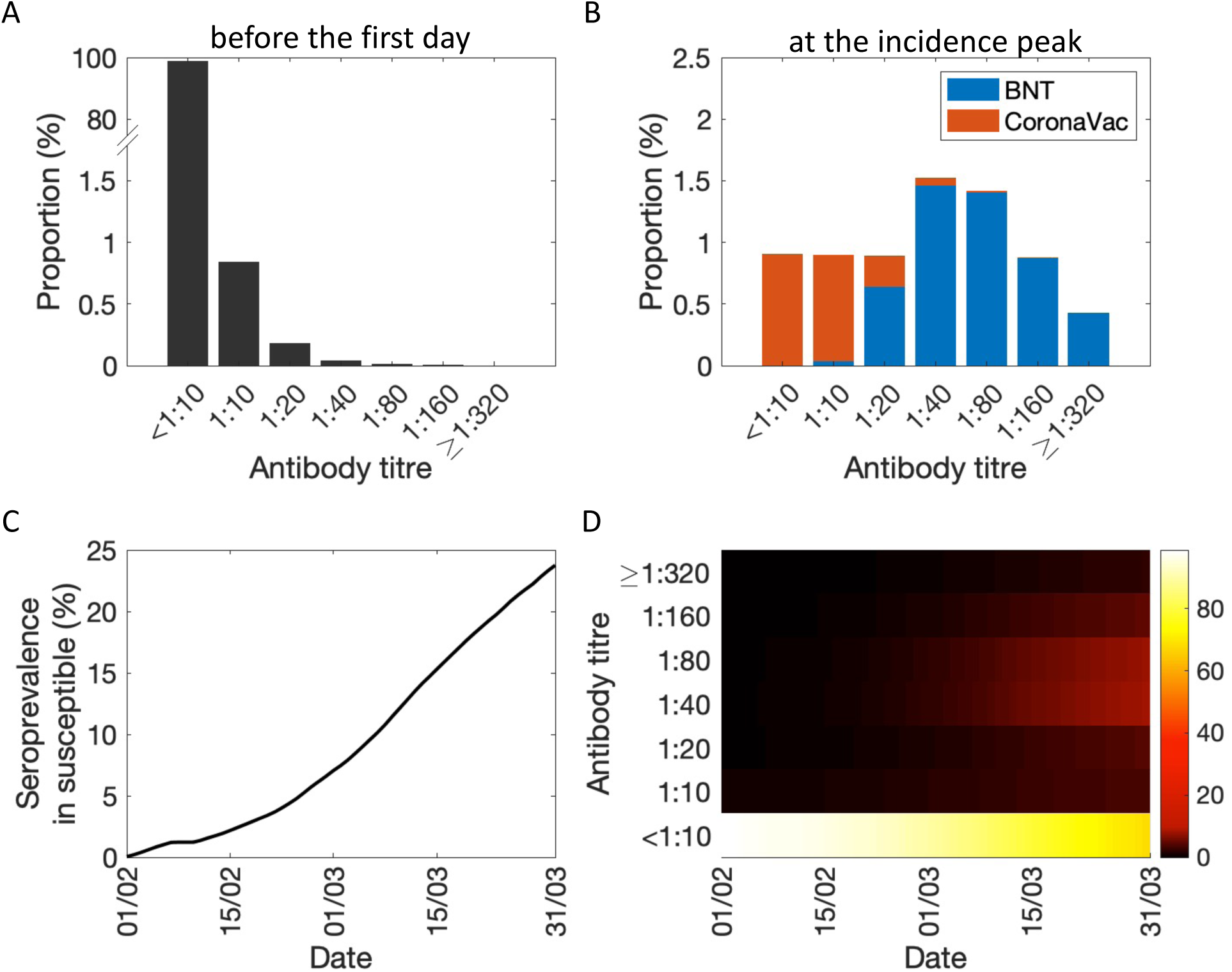
Population immunity and the associated protectiveness. (A) The estimated pre-existing immunity (i.e. the distribution of antibody titers among the population) on 1 February, 2022. (B) Vaccine-induced population immunity at the predicted peak time of the fifth wave (23 February). (C) Seroprevalence (i.e. the percentage of people whose antibody titre ≥1:40) in susceptible individuals. (D) Changes in the proportion of susceptible individuals with each antibody titre level over time.

With the rapid spreading of the omicron virus, many individuals who obtained two doses have continued to take the vaccine booster. According to the daily booster rate (Figure 1C), about 9.9% of the population have taken the booster doses (512,321 for BioNTech and 226,920 for CoronaVac) until one week before the estimated incidence peak time (23 February), and only 4.3% of the population were estimated to have antibody titre ≥1:40, defined as seroprevalence (Figure 3B). The immunological dynamics show that the predicted seroprevalence in susceptible individuals due to vaccination is relatively low (Figure 3CD). Up to the end of February, vaccine booster enabled about 7.5% reduction in the force of infection (Figure S5). Hence, the reduction of *Re* during the late February was difficult to explain by vaccine booster as the vaccine-induced population immunity was still very low.

### Impacts of individual interventions and weather factors

Next, we assessed the impact of each significant NPI (see Table 1). Social distancing regulations were tightened three times. The first tightening (T1) was maintained from 7 January until 10 February with the population mobility reduced by 26.4% on average since 1 February compared to the population mobility before the pandemic began in 2020. The second tightening (T2) was introduced on 10 February up to 23 February with the mobility further to be reduced by 31.1% on average compared to the baseline level. The third tightening (T3) was introduced on 24 February, which allowed the population mobility reduced by 36.4% on average until 31 March. In addition to PCR test, since 26 February, RAT was allowed to be used for confirming infection.

**Table 1.** Description of significant nonpharmaceutical interventions and their impacts on the infections. Predicted number of cumulative incidence resulting from T1 or T2 was calculated assuming that after the end of each tightening, the mobility still maintained at the average level during its implemented period.

| NPIs | Description | Predicted cumulative infections (%) | Effects (percent reduction in cumulative infections) |
| --- | --- | --- | --- |
| First social distancing tightening (T1) | From 7 January, the Government tightened social distancing measures to the same level as used at the epidemic peak time in the wave of the epidemic. A person must wear a mask all the time while on public transportation or in a specified public place [33].<br>Mobility reduced to -26.4% on average between 01 – 09 February. | 58.2 (54.2 – 61.5) | – |
| Second tightening (T2; from 10 February to 23 February) | From 10 February, the maximum number of people permitted for gatherings in public places was reduced from 4 to 2. The maximum number of persons per table in catering premises was 2 except for people presenting their vaccination records in certain premises [34].<br>Mobility reduced to -31.1% on average between 01 – 23 February. | 49.1 (45.2 – 52.7) | 15.6 (14.1 – 17.1) |
| Social distancing further tightening (T3; from 24 February) | Starting on 24 February 2022, all persons shall wear a mask in any public places. The maximum number of persons per table in catering premises was reduced to 2 [35].<br>Mobility further reduced to -36.4% on average between 24 February and 31 March. | 44.5 (40.5–48.1) | 9.5 (8.7 – 10.5)<br>(23.6 (21.8 – 25.6) if compared to T1) |
| Adopting rapid antigen test | Starting on 26 February, members of the public tested positive by RAT, whether distributed by the Government or on their own purchase, should be considered positive cases and they should take all necessary steps to avoid further spreading of the virus, including staying at home [4].<br>Distributing rapid test kits based on the risk levels of districts instead of issuing compulsory testing notices [5]. | 39.0 (36.7–41.1) | 13.2 (11.4 – 15.5) |

Model predicted that, among all major interventions, if only T1 was used, the proportion of cumulative infections increased from 39.0% to 58.2% (95%CI: 54.2-61.5%) (Figure 4A). The subsequent implementation of T2 and T3 further reduced the proportion of cumulative infections to 49.1% (95%CI: 45.2-52.7%) and 44.5% (95%CI: 40.5-48.1%), respectively. The model estimated that about 19.1% (95%CI: 17.7-20.5%) of infectious cases were detected by RAT. With the use of RAT, the proportion of cumulative infections further decreased to the estimated proportion.

**Figure 4.**
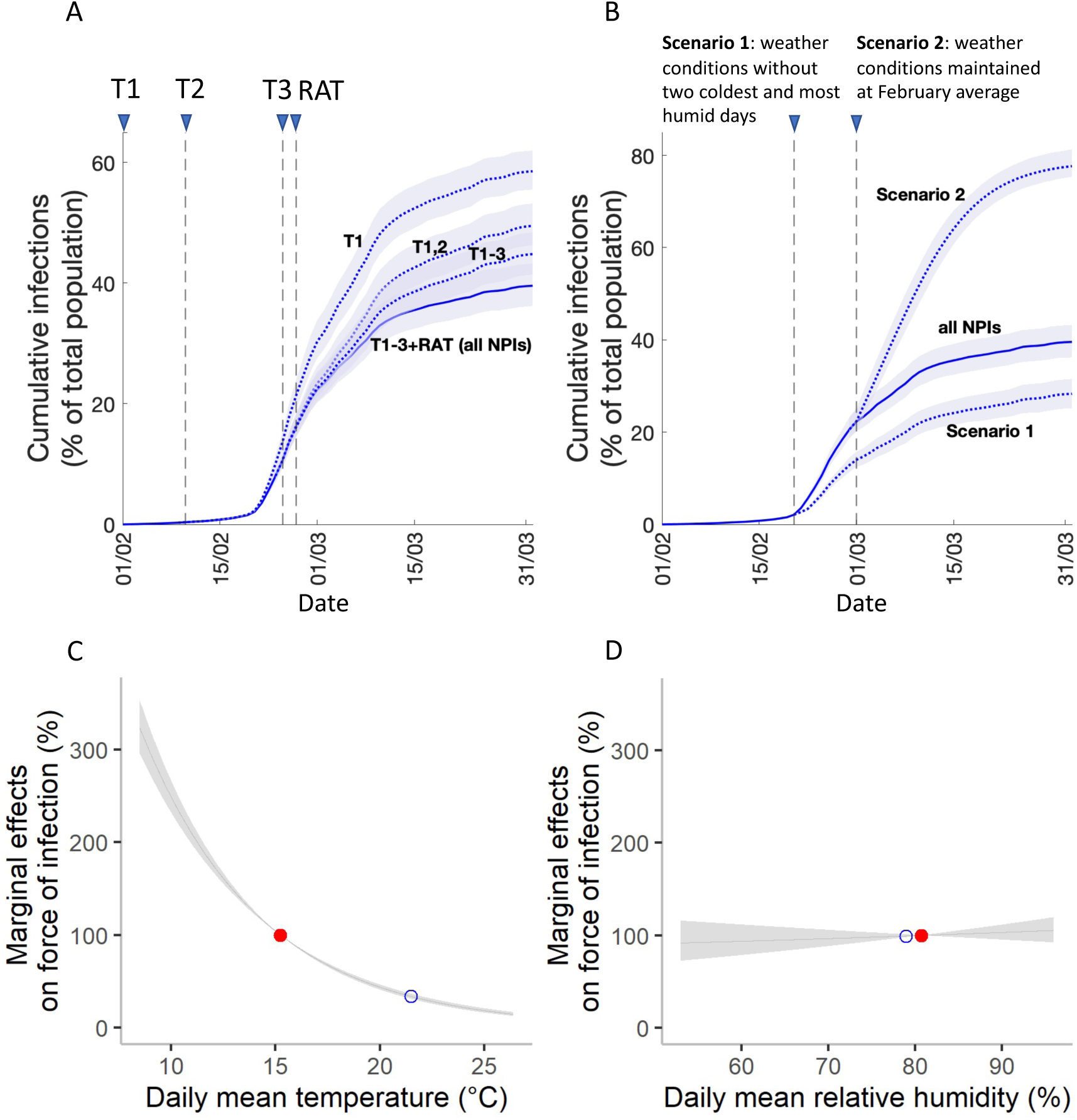
Predicted impacts of individual non-pharmaceutical interventions (NPIs) or changes in weather conditions on cumulative infections. (A) The predicted cumulative infections are plotted in the solid line (with all NPIs) and in the dotted lines (with different combinations of interventions). The triangles represent different start date of the interventions. (B) The predicted cumulative infections of all NPIs under different assumptions of weather conditions. Scenario 1: the coldest and most humidity days (20 and 21 February) are replaced by the average February daily mean temperature and mean relative humidity. Scenario 2: the weather conditions in March are replaced by the average February weather level. (C, D) The marginal effects of temperature and humidity on the force of infection. The red solid points represent the effects of average temperature (C) and average relative humidity (D) in February, and the blue circles represent the average temperature (C) and relative humidity (D) in March.

On the other hand, temperature was found to be associated with the force of infection significantly. The model estimated that an increase of 1°C was associated with a 16.0% (95%CI 14.9 – 17.1) relative reduction in the force of infection and therefore *Re* (see Figure 4C). The average increase of temperature from 15°C to 22°C between February and March (Figure 1C) was thus associated with 66% reduction in the transmissibility. One percent increase in relative humidity was associated with only a 0.3% relative increase in the force of infection (Figure 4D). The average reduction of relative humidity from 81% (February) to 77% (March) was thus associated with about only 1% reduction in the transmissibility.

We further projected the total number of infections under different scenarios of weather conditions. Assuming that the relationship between weather conditions and force of infection held, if the coldest (8.5–10.7°C) and most humid (94–95% relative humidity) days (20 and 21 February 2022) were replaced by the average February daily mean temperature and relative humidity, the predicted cumulative infections reduced significantly to 28.2% (95%CI 25.0–31.5) (Figure 4B). If the weather in March were still maintained as the average February’s level, cumulative infections increased to 77.5% (95%CI 75.1–81.1) up to the end of March (Figure 4B), which is similar as the prediction from the reduced model without weather effects (Figure 2F).

In order to verify whether the sharp pattern of *Re* was affected by NPIs or vaccination, we further simulated *Re* after removing T2&T3, T3, RAT or vaccine boosters. We found that *Re* was generally similar with a moderate level of upward shift (Figure S6). To validate that weather was a key driver of the transmission, we simulated using the reduced model without weather effects. The peak of *Re* was then disappeared (Figure 2B).

### Required interventions for outbreak prevention

Our simulation results showed that the COVID-19 transmission was not easy to be managed by social distancing, RAT and pre-existing vaccine booster in a cooler condition. For example, even with 80% booster coverage, mobility needs to be reduced at least 65% (Figure 5A), which was far higher than the actual average reduction in mobility observed during a more relaxed T1 period (26.4%) or a more tightened T3 period (36.4 %). Otherwise, RAT coverage (i.e. the percentage of cases that are detected by RAT) has to be greater than 60% during the more relaxed T1 period (Figure 5C).

**Figure 5.**
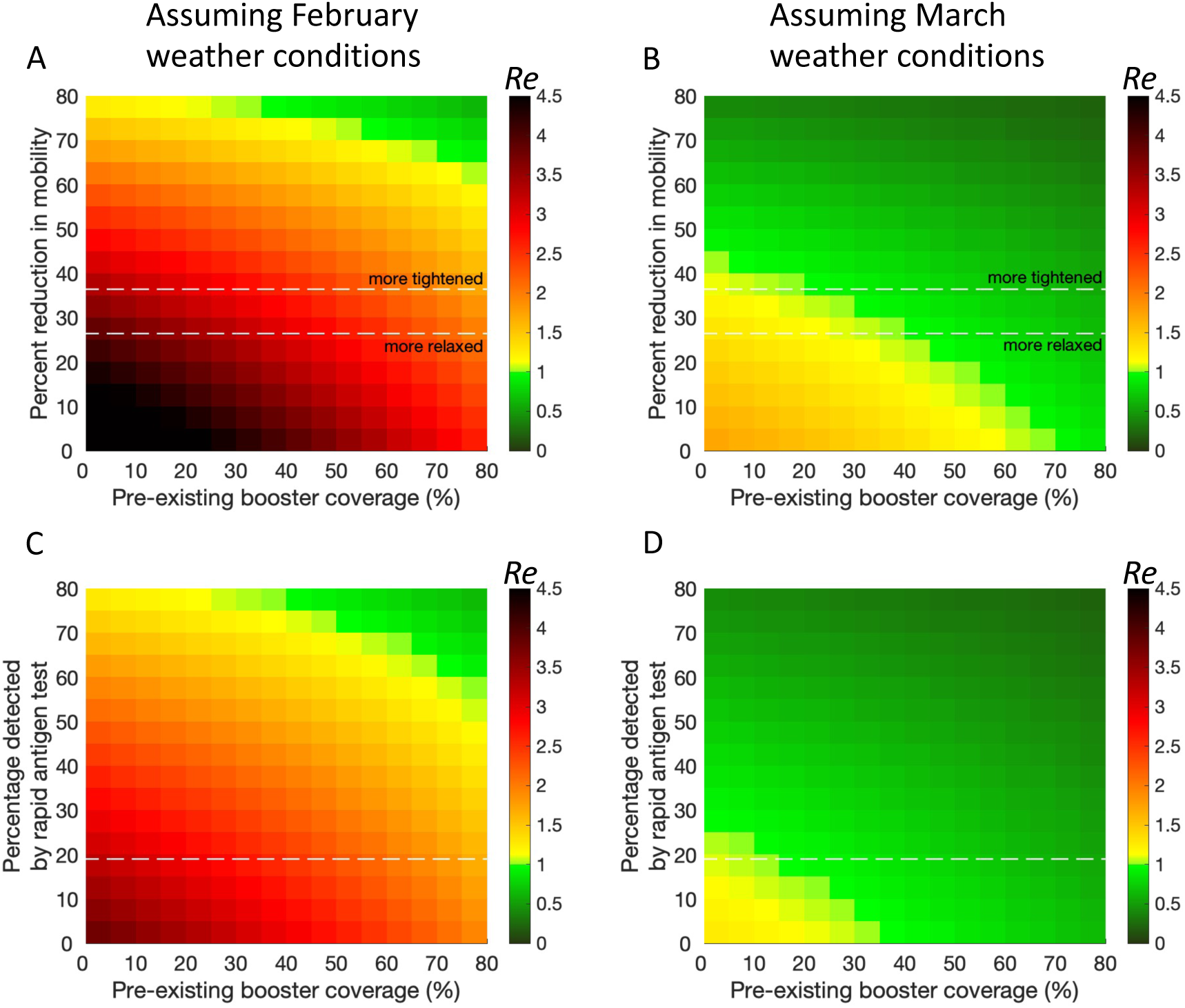
The impacts of mobility and rapid antigen test on transmissibility with different pre-existing booster coverages and different weather conditions. (A) *Re* plotted for a combination of percent reduction in mobility and booster coverage with an average weather condition in February without the uses of RAT. The upper dashd line (labeled as more tightened) represents the average mobility level when T3 was used. The lower dashed line (labeled as more relaxed) represents the average mobility level between 1 and 9 February, when T1 was used. (C) *Re* plotted for a combination of different coverage of RAT and vaccine booster with an average weather condition in February. T1 was used. The dashed line represents the model-estimated percentage of cases that are detected by RAT (19.1%). The second column, (B) and (D), shows the same data as (A) and (C) but with an average weather condition in March. In (A) and (B), the RAT coverage is set as 0. In (C) and (D), mobility is fixed as the average level during T1, as the more relaxed tightening.

In comparison, if the average weather conditions in March were assumed, *Re* of below 1 can be achieved without the uses of RAT when about one third of people had taken booster doses during the more tightened period (Figure 5B), meanwhile, without a surge on *Re*. 10% RAT coverage allowed social distancing measures to relax as T1 (Figure 5D).

## Discussion

In countries and cities that have adopted a zero-Covid policy [3] [25], their low population immunity indicates high risks of infection and death. This study used Hong Kong, a densely populated city in southern China with low pre-existing booster coverage [6] as an example to assess the impacts of interventions and seasonal factors (temperature and humidity).

Modelling results suggest that cold temperatures were associated with the sharp increase in transmissibility during the early Hong Kong’s fifth waves (Figure 1C, 2C and 4C). Although social distancing tightenings and vaccine booster posed important impacts on mitigation, temperatures were more likely to be the main factor to drive or contain the spread of the outbreak (Figure 4B, Figure S1). These results were validated after comparing the full model to the model without weather effects (Figure 1). On the other hand, a very weak association between relative humidity and *Re* was found. The relationship between relative humidity and the transmissibility has been studied but with different conclusions [12][13][14][26].

There are some explanations of the impact of lower temperature on disease transmission. First, the virus can be more stable at cold condition, which facilitates its transmission [27] [28]. Second, temperature may affect the immune responses of hosts, or moderate the interaction between the virus and host immune system [29]. Third, an alternative explanation is that people may spend more time indoors in cold days, which may increase the likelihood of indoor airborne transmission [30]. Possible mechanisms for temperature-dependent infectivity in SARS-CoV-2 has been discussed [10].

### Policy Relevance

We suggest that temperature pattern should be taken into account for making public health decisions, such as deciding the level of NPIs or the timing of booster doses. Our results (Figure 5AC) found that, if the local spread begins during a cold weather condition, the outbreak is likely to be extremely difficult to suppress (Re <1). Very high booster coverage (around 80%) combined with extremely strengthed social distancing (stronger than all tightenings introduced in the fifth wave) or frequent uses of RAT (e.g. more than half of the cases are detected by these kits) are required. If contact tracing cannot prevent initial transmission clusters from growing, high vaccine booster coverage with mass testing or regional lockdown seems to be unavoidable to suppress the Omicron outbreak. On the other hand, the outbreak is likely to become more manageable during warmer seasons.

Additional doses or boosters are likely to be a standard part of the vaccination schedule, similar as the scenario seen for seasonal influenza [31]. Because of antibody waning, getting vaccinated too early before the next outbreak may result in insuffcient immunity when infections occur. As the current pandemic is likely to become seasonal epidemics [32], having the additional doses at a ‘right’ timing (e.g. before winter) may be another important factor, in addition to the vaccine coverage, for reducing the total incidence.

These findings are not implying that public health interventions are not important during warmer seasons. We suggest that seasonal temperature variation should be taken into account for policy making or model projection. For example, up to now (the middle of May 2022), even though social distancing measures have been relaxed more than previous T1 (mobility also increases to the level before the fifth wave) together with about only 50% booster coverage, cases are very limited (< 200 per day), which is consistent to our projection assuming warm conditions (Figure 5D).

### Limitations

Some limitations exist in our study. First, the study mainly focus on the disease transmissibility while not explore the impact of vaccine or seasonal factors on disease severity. Second, the number of total infections may be underestimated since the proportion of cases that are underreported is largely unknown when the testing capacity is limited. Therefore, we attempted to capture the changes in underreporting and reporting delay in our modelling. Third, model validation may be sensitive to the assumption of the protectiveness of vaccine or natural infections. Here the data used in our study were based on a published empirical study without age stratification [7].

### Conclusion

A recent work has suggested a striking effect of temperature on the spread of COVID-19 [10]. Here, we found that temperature was associated with a larger impact on the transmissibility than strict public health interventions without lockdown throughout a significant outbreak in a single city. Incorporating seasonal variation in temperature can improve the accuracy of modelling of SARS-CoV-2 transmission, which helps to find a balance between normal life and low health impact.

## Data Availability

All data produced in the present study are available upon reasonable request to the authors.

