## Supplementary Materials for "Modelling the impacts of public health interventions and weather on SARS-CoV-2 Omicron outbreak in Hong Kong"

Hsiang-Yu Yuan[a,b] and Jingbo Liang[b]
[a]Centre for Applied One Health Research and Policy Advice, Jockey Club College of Veterinary Medicine and Life Sciences, City University of Hong Kong, Hong Kong SAR, China
[b]Department of Biomedical Sciences, Jockey Club College of Veterinary Medicine and Life Sciences, City University of Hong Kong, Hong Kong SAR, China

#### Supplementary Figures

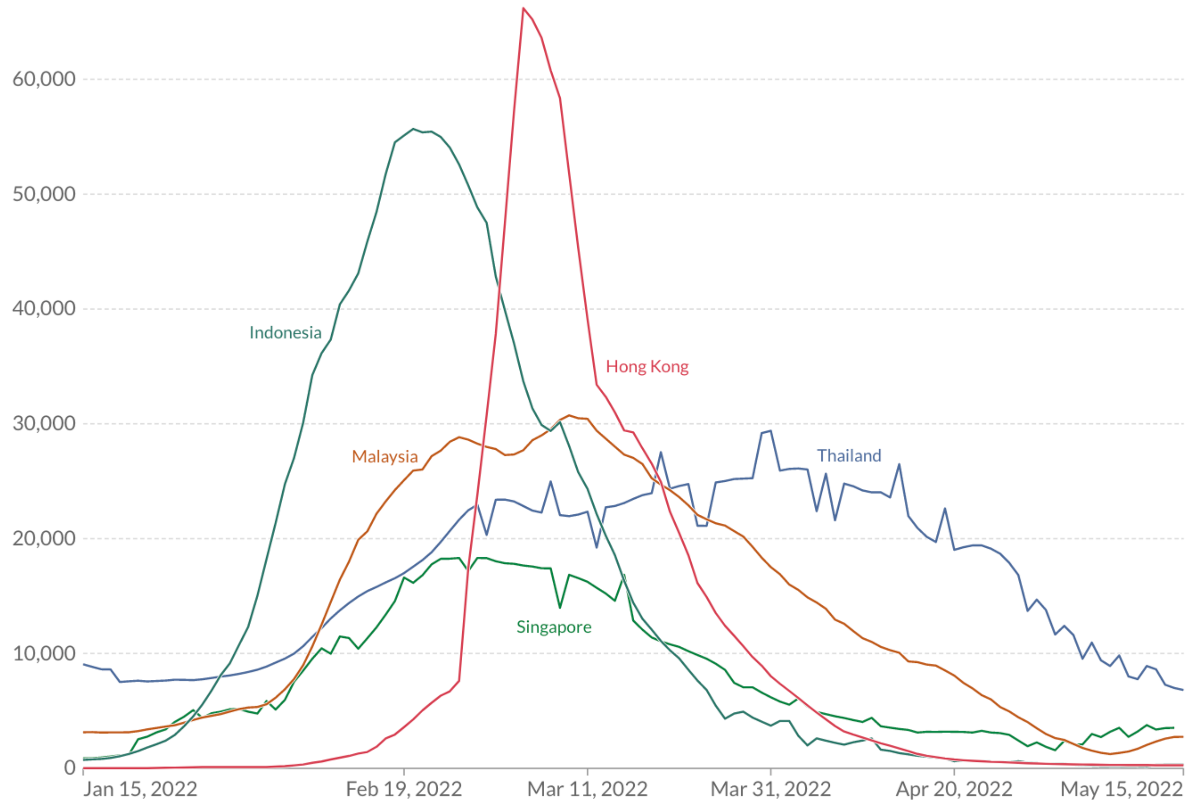


Figure S1. Comparison of the outbreaks between Hong Kong and nearby tropical countries occurring during the same period; Picture adopted from Our World in Data [1]. For comparison, except Hong Kong, only countries whose major cities are below 20 degrees north latitude and daily case number increases above 10,000 after January are included. Source: Johns Hopkins University CSSE COVID-19 Data.

*Vaccination, NPI, and reporting*

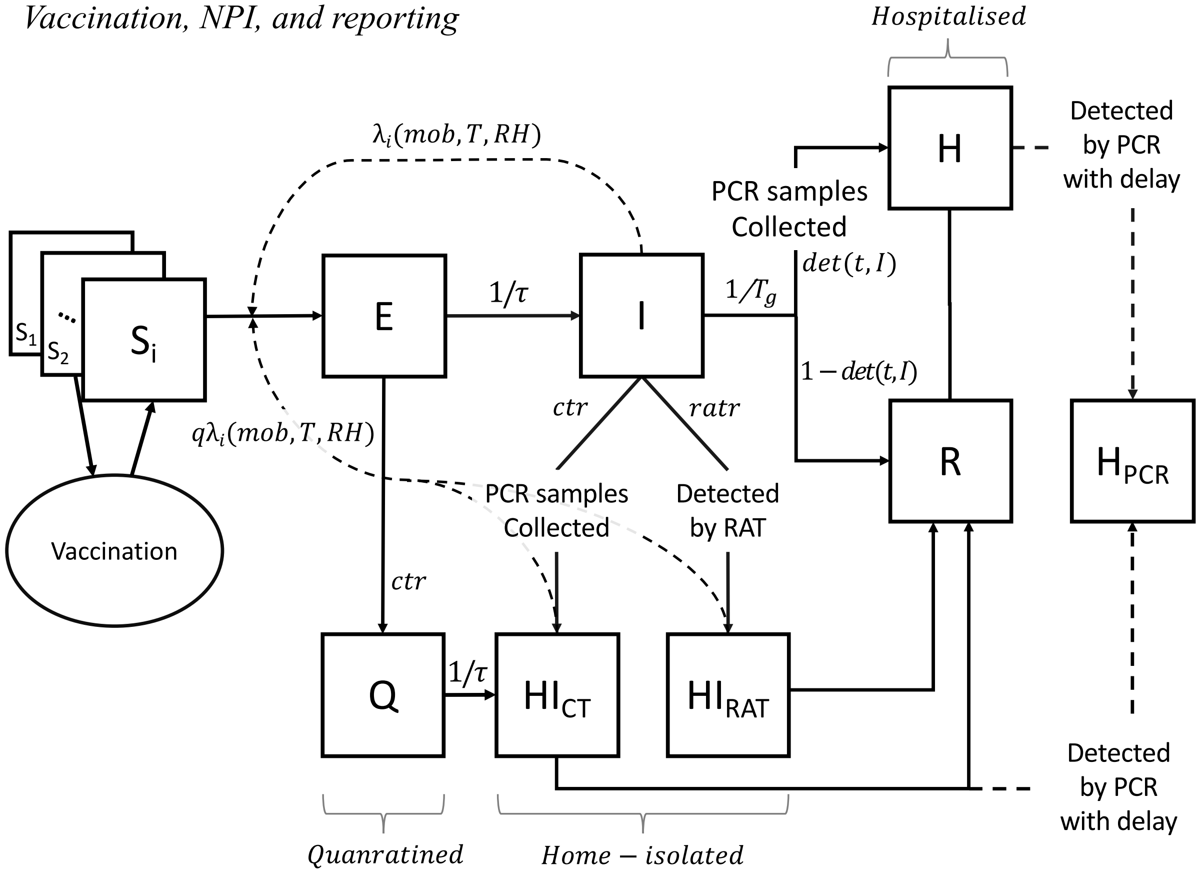


Figure S2. Schematic of Susceptible (*S*), Exposed (*E*), Infectious (*I*), Recovered (*R*) model with vaccination, non-pharmaceutical interventions and dynamics reporting rates. *Si* represents the susceptible individuals with *i*th antibody level. *Vaccination* represents the change of antibody titre levels. The force of infection of *Si* is denoted as $\lambda i$, which is a function of daily mobility (*mob*), temperature (*T*) and relative humidity (*RH*). *q* is a factor to reduce the force of infection among home-isolated cases. Cases detected by PCR includes hospitalised cases (*H*) and home-isolated cases that are contact-traced ($HI_{CT}$). Cases detected by RAT refer to home-isolated cases after self-testing ($HI_{RAT}$). *det*, *ratr* and *ctr* and refer to the detection rate by PCR, the detection rate by RAT and the rate of being contact-traced. Some of the cases are traced and quarantined before being infectious (*Q*). $H_{PCR}$ represents the daily cumulative number of cases detected by PCR with testing delay.

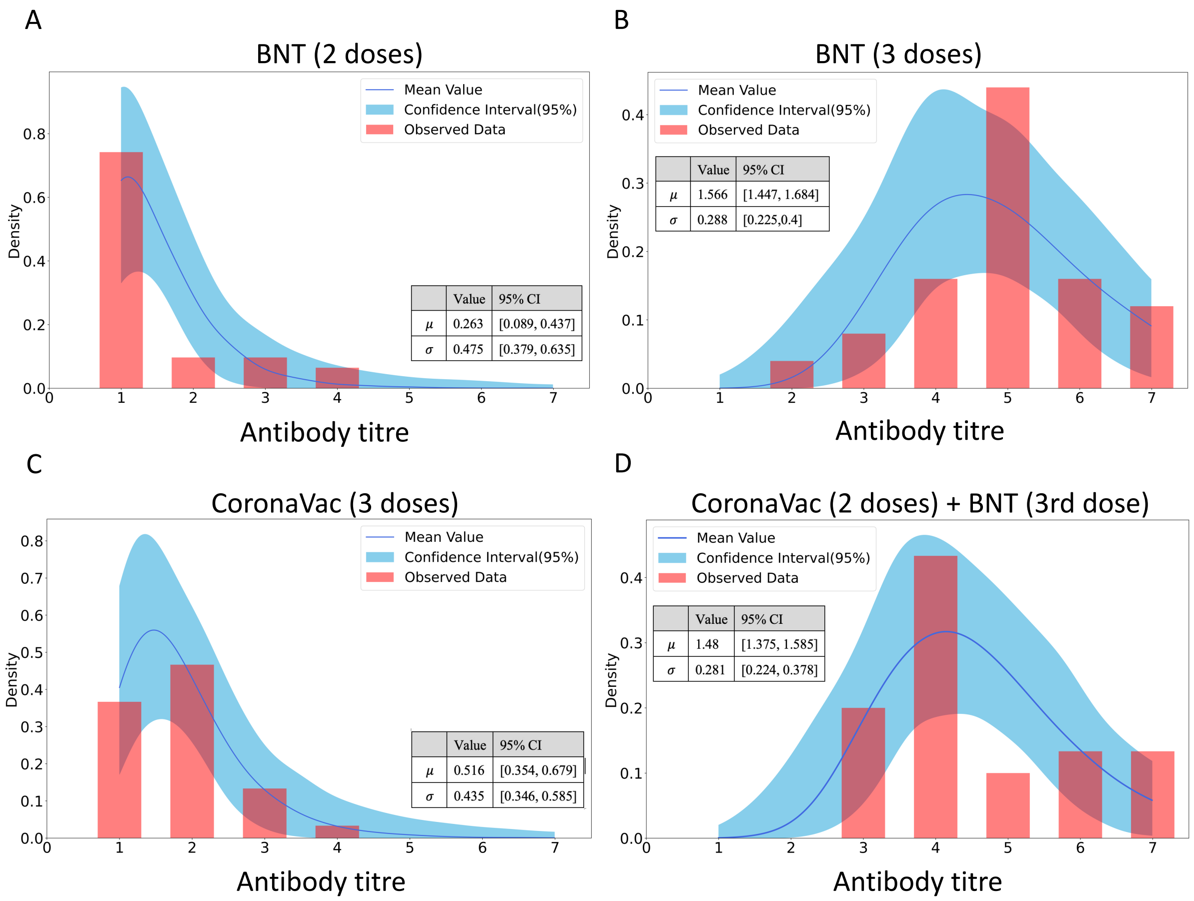


Figure S3. The distribution of antibody responses after vaccination. (A) Antibody titres after full immunisation of BNT. (B) Antibody titres after BNT booster. (C) Antibody titres after CoronaVac booster. (D) Antibody titres after full immunisation of CoronaVac and one booster dose of BNT.

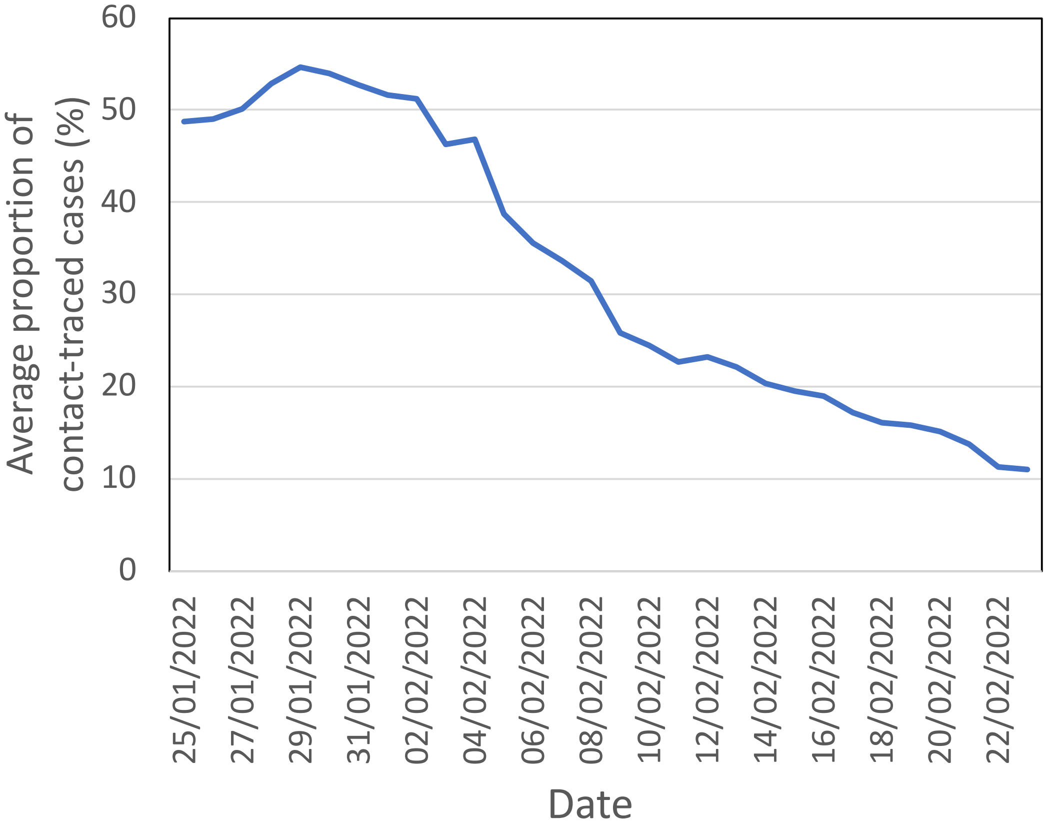


Figure S4. The average proportion of cases that are contact-traced in the past four weeks. The average proportion was calculated using the data only considering local cases collected from the Hong Kong Centre for Health Protection [2]. Note that data after 22 February, 2022 were not available.

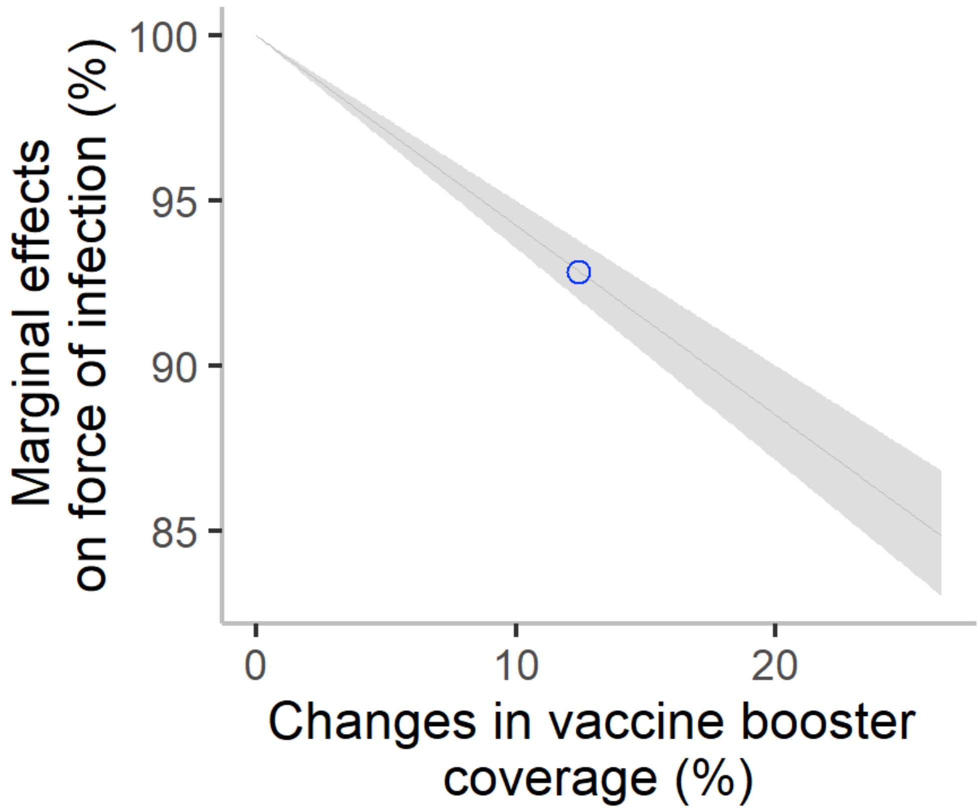


Figure S5. The marginal effects of vaccine booster coverage on the force of infection. Blue circle represents the increase of vaccine booster coverage during February, 2022.

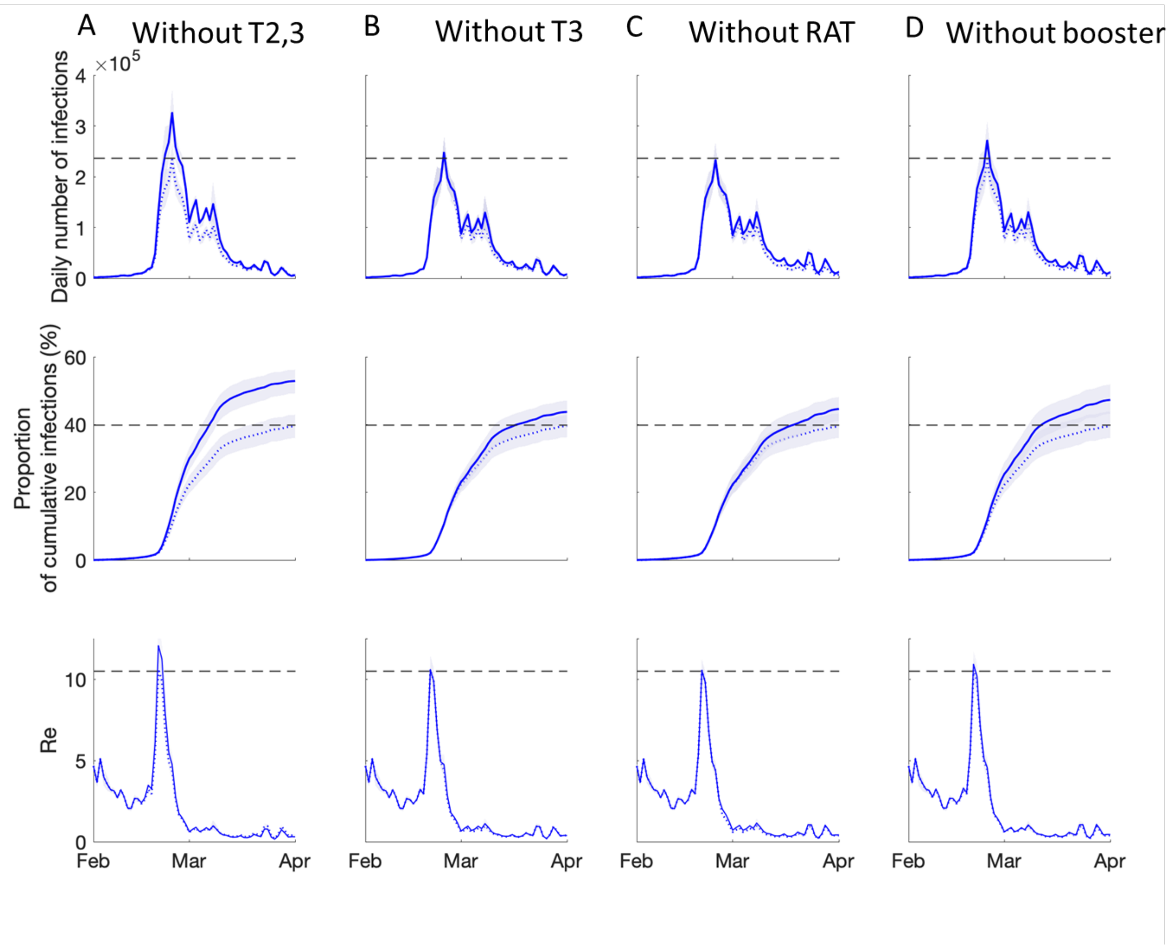


Figure S6. Impacts of social distance tightenings, RAT, and vaccine booster on transmission dynamics and transmissibility of the fifth wave. (A) Daily number of infections, proportion of cumulative infections and Re when the full interventions were implemented except T2 and T3. (B,C,D) represent full interventions without T3 (B), RAT (C), or vaccine booster administration (D). The solid lines are the infection and transmissibility dynamics simulated from different combinations of interventions, while the dotted lines represent the dynamics when the full interventions were implemented.

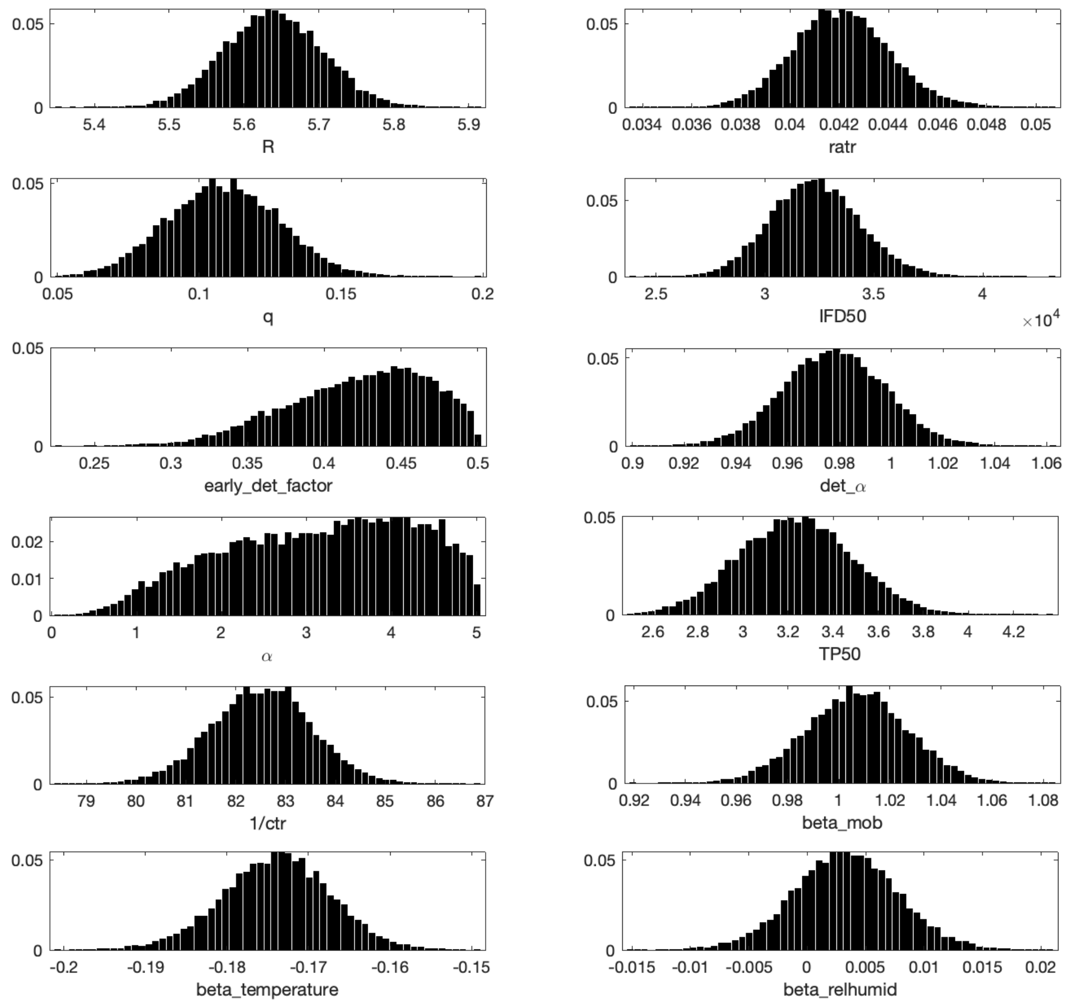


Figure S7. Posterior distributions of epidemiological parameters and the parameters of NPIs, vaccine protectiveness, and weather conditions.

### Supplementary Tables

Table S1. Definitions and posterior estimates of the parameters of NPIs, vaccination protectiveness, and weather conditions. Mean values with 95% credible intervals are produced after $10^6$ McMC steps.

| **Parameter** | **Symbol** | **Value (95% CI)** |
|---|---|---|
| Baseline reproduction number | *R* | 5.64 (5.51 - 5.77) |
| The rate of RAT uses | *ratr* | 0.042 (0.038 - 0.046) |
| Inverse of contact-tracing rate | *1/ctr* | 1.90 (1.72-2.12) |
| Ratio of the contact rates of quarantined to unquarantined individuals | *q* | 0.109 (0.071 – 0.147) |
| Threshold number of infectious cases that enables 50% to be detected | *IFD50* | 32,313 (28,484 – 36,475) |
| Scaling factor to adjust detection rate after reporting flow was simplified | *early_det_factor* | 0.423 (0.326 – 0.493) |
| Shape parameter to adjust early_det_factor | $det\_\alpha$ | 0.978 (0.939 – 1.017) |
| Shape parameter of the sigmoid function | $\alpha$ | 3.149 (1.012 – 4.874) |
| Titre level associated with 50% protection | *TP50* | 3.234 (2.756 – 3.717) |
| Regression coefficient of mobility reduction | $\beta_{MOB}$ | 1.007 (0.097 – 1.046) |
| Regression coefficient of temperature | $\beta_T$ | -0.174 (-0.187 – -0.161) |
| Regression coefficient of relative humidity | $\beta_{RH}$ | 0.003 (-0.005 – 0.012) |

Table S2. Antibody titre levels in different cohorts after 2 or 3 doses of BNT and CoronaVac. Data are collated from a previous study [3].

| **Titre level** | **1** | **2** | **3** | **4** | **5** | **6** | **7** |
|---|---|---|---|---|---|---|---|
| BNT162b2(2 doses) | 23 | 3 | 3 | 2 | 0 | 0 | 0 |
| BNT162b2(3 doses) | 0 | 1 | 2 | 4 | 11 | 4 | 3 |
| CoronaVac(3 doses) | 11 | 14 | 4 | 1 | 0 | 0 | 0 |
| CoronaVac(2 doses) + BNT162b2(3rd dose) | 0 | 0 | 6 | 13 | 3 | 4 | 4 |

### Supplementary Methods

We developed a stratified immunity Susceptible-Exposed-Infectious-Recovered model [4] and included NPIs [5] and daily uncertainties (such as vaccination, mobility, temperature and relative humidity). In order to fit the model to observed data, we also introduced a time-varying reporting rate and delay (Figure S2). Sections *Transmission model* and *Fitting antibody responses* are described as below.

#### 1. Transmission model

*A. Model assumptions*

The size of population was set to be 7.48 million people. The following assumptions were made for modelling the infection dynamic of the fifth wave in Hong Kong. We assumed that people became fully protected after natural infection during the study period because we did not observe any report of reinfection event. These recovered people did not take vaccine booster later. Therefore, people took the vaccine booster before being infected or when they had not been infected. Because of the short period, waning immunity after vaccine booster or natural infections was therefore not considered during the study period.

*B. Transmission mechanism with stratified immunity*

The transmission among individuals in each titre level was formulated as

$$\frac{dS_i}{dt} = -S_i\lambda_i + Vaccination(i) \quad (1)$$

$$\frac{dE}{dt} = \sum_{i=1}^{maxi} S_i\lambda_i - (\frac{1}{\tau} + ctr)E \quad (2)$$

$$\frac{dI}{dt} = \frac{1}{\tau}E - (ctr + ratr + \frac{1}{T_g})I \quad (3)$$

$$\frac{dQ}{dt} = ctrE - \frac{1}{\tau}Q \quad (4)$$

$$\frac{dH}{dt} = \frac{det(t,I)}{T_g}I \quad (5)$$

$$\frac{dHI_{ct}}{dt} = ctrI + \frac{1}{\tau}Q - \frac{1}{T_g}HI_{ct} \quad (6)$$

$$\frac{dHI_{rat}}{dt} = ratrI - \frac{1}{T_g}HI_{rat} \quad (7)$$

$$\frac{dH_{pcr}}{dt} = \frac{1}{delay}(H + HI_{ct}) \quad (8)$$

where $S$, $E$, $I$, $Q$, $H$, $HI_{ct}$, $HI_{rat}$, $H_{pcr}$ and $R$ refer to individuals that are susceptible ($S$), exposed ($E$), infectious ($I$), quarantined after being contact-traced during the latent period ($Q$), hospitalised after being detected by PCR ($H$), home-isolated after being contact-traced ($HI_{ct}$), home-isolated after being detected by RAT ($HI_{rat}$), PCR-detected ($H_{pcr}$) and recovered or fully protected ($R$). $\lambda_i$ is the force of infection for $S_i$ whose antibody titre level is $i$; $\frac{1}{\tau}$ is the incubation time (assumed to be 3 days); $T_g$ is the infectious period (assumed to be 6 days); $ctr$ is the rate of being contact-traced; $ratr$ is the detection rate of being tested positive using RAT; and $det(t, I)$ is the detection rate of being tested positive using PCR, which is a function of the number of infectious individuals.

In order to match the antibody boosting experimental results (see section *Fitting antibody responses*), we assume that the minimum titre level is 1 and the maximum titre level is 7 to represent the antibody titres from <1:10, 1:10, …, 1:160 to ≥ 320. The term $Vaccination(i)$ gives the change in the number of susceptible individuals having antibody titre level of $i$ (see the section in *Antibody boosting after vaccination*).

The model is able to produce daily numbers of cases detected by either PCR or RAT. These test methods had different consequences:

1. PCR. Due to the limited capacity of testing, many of the cases that were detected by PCR were more severe. They were hospitalised or stayed at isolation facilities ($H$). Some PCR-detected cases were belonging to possible close contacts of infected cases and were required for compulsory testing. Some of them were traced before symptoms developed and they were detected after latent period ($Q$). If the symptoms were not severe, they self-isolated at home ($HI_{ct}$). Assuming that the testing delay of PCR is $delay$, the cumulative number of reported cases detected by PCR is calculated as $\Delta H_{pcr}(\tau) = \int_{(\tau-1)}^{\tau} \frac{1}{delay}(H + HI_{ct})dt.$

2. RAT. Many cases were detected by themselves using RAT when hospitals were approaching full capacity. These cases self-isolated at home ($HI_{rat}$) and reported their positive results through an official online system without testing delay. Hence, daily number of reported cases detected by RAT is calculated as $\Delta HI_{rat}(\tau) = \int_{(\tau-1)}^{\tau} ratrI\ dt.$

Reporting delay for PCR was assumed to be between 0.1 and 1.2 days after checking the confirmation delay of the reported cases [2]. The number was depending on the number of infectious cases.

$$delay = (T_{max} - T_{min})(1 - e^{-\left(\frac{\mathrm{Iprev}-30000}{300000}\right)}) + T_{min} \qquad (9)$$

where $T_{min}$= 0.1 and $T_{max}$= 1.2 days.

The force of infection $\lambda_i$, the rate at which susceptible individuals having antibody titre level i become infected is calculated as follows:

$$\lambda_i = susc_i \cdot \frac{R_0}{T_g} \cdot (1 + \beta_m mob(t)) \cdot e^{\beta_T(T(t)-T_0)} e^{\beta_{RH}(RH(t)-RH_0)} \cdot \left[\frac{I + q(HI_{rat} + HI_{ct})}{N}\right] \qquad (10)$$

where the term $susc_i \cdot \frac{R_0}{T_g} \cdot (1 + \beta_m mob(t))$ represents the transmission rate of an infectious individual who are not under quarantine or home-isolation. $susc_i$ is the relative susceptibility of infection for susceptible individuals having antibody titre level $i$. $mob(t)$ is the change of population mobility index compared to the reference level at pre-pandemic period [6]. $\beta_m$ is the coefficient for $mob$. $\beta_m mob(t)$ measures the degree of social mixing at time $t$. The term $e^{\beta_T(T-T_0)}$ represents the effect of temperature [7], where $T(t)$ is the daily temperature and $T_0$ is the baseline temperature (i.e. the average temperature in February). $\beta_T$ is the coefficient for temperature. $\beta_{RH}$ is the coefficient for relative humidity. $RH(t)$ relative humidity and $RH_0$ is the baseline relative humidity (i.e. the average relative humidity in February). Individuals can also be infected by infectious cases isolated at home. The term $HI_{rat} + HI_{ct}$ represents the number of home-isolated cases (who are still infectious) and $q$ is a scaling factor representing a reduction of social interactions between home-isolated cases and others [5]. $N$ is the population in Hong Kong.

Following our previous study [4], the unnormalized susceptibility of individual having antibody titre level was defined as:

$$susc_i^{un} = \frac{1}{1+e^{\alpha(i-TP50)}} \tag{11}$$

where $i$ (i.e. 1 to 7) represents the antibody level from <1:10, 1:10, 1:20,…, 1:160, and ≥1:320 [3]; and $TP50$ represents a threshold level which gives 50% protection. Because 1:25.6 (95%CI: 18.3–36.0) was found to exhibit 50% protection [3], $TP50$ was calculated as log(25.6/5)+1, which was 3.36 (95%CI: 2.87-3.85). α was a shape parameter that controls the curvature of the function of susceptibility. $susc_i$ is the normalized susceptibility that allows maximum susceptibility to be 100%.

*C. Time-varying detection rate*

We assumed that generally the PCR detection rate $det(t)$ becomes lower when the actual number of infectious cases without being quarantined or home-isolated was higher. The baseline detection rate is:

$$det_{base}(I) = (k_{max} - k_{min})e^{-\log(2)\left(\frac{I(t)}{IFD50}\right)} + k_{min} \tag{12}$$

This allows the detection rate declines from maximum PCR detection rate $k_{max}$ (set to be 80%) to minimum $k_{min}$ (set to be 10%) as $I(t)$ increases. When $I(t)$ reaches $IFD50$, 50% of infectious cases will be detected. Hence, we called this threshold the number of infectious cases that enables 50% to be detected ($IFD50$). This number was estimated through McMC.

During the early increasing period until the incidence peak has been passed, the detection rate was assumed to be temporarily increased since 26 February because the follow-up procedures for confirming positive results in PCR and RAT tests were streamlined, in order to reduce the number of cases waiting for PCR. Between 26 February and 2 March, the PCR detection rate becomes $det^{before}$:

$$det_{tar}(I) = det_{base}(I) + (1 - det_{base}(I)) \cdot early\_det\_factor \tag{13}$$

$$det^{before}(t, I) = det_{tar}(I) + (det_{base}(I) - det_{tar}(I))e^{-det\_\alpha(t-26)} \tag{14}$$

where $det\_boost\_factor$ is the scaling factor to adjust detection rate after reporting flow was simplified and $det\_\alpha$ incorporates the delay effects of $early\_det\_factor$ to allow a gradual change. After 2 March,

$$det^{after}(t, I) = det_{base}(I) + (det_{tar}(I) - \det(I))e^{-det\_\alpha(t-26)} \tag{15}$$

Therefore, the detection rate will be:

$det(t, I) = det_{base}(I)$ when $t$ is before 26 February
$det(t, I) = det^{before}(t, I)$ when $t$ is between 26 February and 2 March
$det(t, I) = det^{before}(t, I)$ when $t$ is after 2 March

*D. Antibody boosting after vaccination*

The function $Vaccination(i, t)$ represents the change of the number of susceptible individuals having antibody titre level $i$ among people at time $t$. The function includes both the numbers of people removed from and people whose antibody boosted to titre level $i$ after having vaccinated seven days ago, such as:

$$Vaccintation(i, t) = -removed(i, t - 7) + boosted(i', i, t - 7) \quad (16)$$

$$removed(i, t - 7) = dailyvac(t - 7) \times S_i \quad (17)$$

$$boosting(i', i, t - 7) = \sum_{i'=1}^{i} dailyvac(t - 7) \times S_i \times g(i', i) \quad (18)$$

where $vac(t - 7)$ represents the daily vaccination rate 7 days ago before antibody was boosted. We assumed that the effect of vaccination occurs after 7 days. $g(i', i)$ is a lognormal function representing the distribution of antibody titre boosted from level $i'$to $i$. We assumed that antibody titre level only increases or maintain at the same level after vaccination. See the section *Fitting antibody responses* for the parameter estimation of the boosting function. Since antibody response of natural infection by Omicron is largely unknown, for simplicity, we assumed that natural infection increases antibody titre to the highest level, indicating that no reinfection occurs within this outbreak.

*E. Parameter estimation using Markov chain Monte Carlo*

The number of initial infectious cases was set to 2200 before 1 February. The number was determined assuming 80% of these cases were detected and reported in the next 7 days. The number of confirmed cases between 1 February and 7 February was 1760. Hence, we the number of initial infectious cases was obtained. Infectious period was set to be 6 days and incubation time was 3 days.

Likelihood of observing the number of confirmed cases was calculated using a negative binomial with the expected mean value as the number of model-predicted reported cases $H$ at day $t$. The dispersion parameter $r$ was set to be 16.

$$Y(t) \sim NegBinom(\mu(t), r) \quad (19)$$

where $\mu(t)$ is the daily number of newly reported cases. Before the RAT was adopted by government on 26 February, the model was compared only to PCR detected cases, such as $\Delta H_{pcr}(t)$. Since then, the model produced two daily numbers of cases, $\mu 1(t) = \Delta H_{pcr}(t)$ and $\mu 2(t) = \Delta HI_{rat}$.The joint likelihood of these two events was calculated.

The posterior distributions of the parameters of the model were obtained after fitting the model to the daily number of reported cases. The posterior distributions were estimated using a Markov chain Monte Carlo (MCMC) algorithm with at least $10^6$ steps until effective sample size (ESS) of is greater than 1000 for all parameters. Gelman-Rubin convergence diagnostic was calculated for each parameters using the

R package stableGR. All resulting numbers were larger than 1 but less than 1.0006, indicating good convergence.

*F. The reduced model*

A reduced or null model was fit to the same data where the effects of temperature and relative humidity are zero. This is equivalent to allow weather conditions to be constrained to be constant. The whole procedure for data fitting is still same as that for the full model. We were asking whether the effects of weather conditions restrict the epidemic shape, hence affecting model fitting.

### 2. Fitting antibody responses

We assumed that the antibody boosting for an individual taking booster dose follows a log-normal distribution with a mean of $\mu$ and a standard deviation of $\sigma$.

$$g(i', i \mid \mu, \sigma) = \frac{1}{\sqrt{2\pi} i\, \sigma} e^{-\frac{(\ln i - \mu)^2}{2\sigma^2}} \tag{20}$$

where $i$ is a boosted titre level and $i'$ is the original titre. The criteria is the boosted titre ($i$) is greater than or equal to the original titre ($i'$). $\mu$ represents the average increase and $\sigma$ is the standard deviation of the antibody responses. These two parameters were estimated through Maximum Likelihood Estimation (MLE) approach.

*A. MLE for the log-normal distribution*

Now, we have the observed titre levels in different cohorts. Let $X_1, \ldots, X_n \sim$ Lognormal$(\mu, \sigma^2)$.

The likelihood is:

$$L(\mu, \sigma^2) \propto \prod_{i=1}^{n} x_i^{-1} \exp\left[\sum_{i=1}^{n} \frac{-(\ln x_i - \mu)^2}{2\sigma^2}\right] \tag{21}$$

The log-likelihood is:

$$l(\mu, \sigma^2) \propto -\frac{n}{2} \ln \sigma^2 - \frac{1}{2\sigma^2} \sum_{i=1}^{n} (\ln x_i - \mu)^2 - \sum_{i=1}^{n} \ln x_i \tag{22}$$

We can estimate these two parameters by minimizing negative log likelihood:

$$\hat{\mu} = \frac{\sum_{i=1}^{n} \ln x}{n}$$

$$\hat{\sigma}^2 = \frac{\sum_{i=1}^{n} (\ln x - \hat{\mu})^2}{n}$$

*B. Parameter estimates for different vaccination cohorts*

Using the MLE, we obtained the fitted log-normal model in four cohorts. Table S2 shows the titre levels in different cohorts. Parameter estimates of the antibody responses after full immunisation of BNT, after BNT booster, after CoronaVac booster, and after full immunisation of CoronaVac and one booster dose of BNT are shown in Figure S3.

### References


1 Our World in Data. Coronavirus (COVID-19) Cases. https://ourworldindata.org/covid-cases

2 Hong Kong Centre for Health Protection. https://www.chp.gov.hk/en/index.html (accessed 1 May 2022).

3 Cheng SMS, Mok CKP, Leung YWY, *et al.* Neutralizing antibodies against the SARS-CoV-2 Omicron variant BA.1 following homologous and heterologous CoronaVac or BNT162b2 vaccination. *Nat Med* 2022;**28**:486–489.

4 Yuan HY, Baguelin M, Kwok KO, *et al.* The impact of stratified immunity on the transmission dynamics of influenza. *Epidemics* 2017;**20**:84–93. doi:10.1016/j.epidem.2017.03.003

5 Yuan HY, Blakemore C. The impact of multiple non-pharmaceutical interventions on controlling COVID-19 outbreak without lockdown in Hong Kong: A modelling study. *Lancet Reg Heal – West Pacific* 2022;**20**:100343. doi:10.1016/J.LANWPC.2021.100343

6 Google. COVID-19 Community Mobility Reports. https://www.google.com/covid19/mobility/

7 Smith TP, Flaxman S, Gallinat AS, *et al.* Temperature and population density influence SARS-CoV-2 transmission in the absence of nonpharmaceutical interventions. *Proc Natl Acad Sci U S A* Published Online First: 2021. doi:10.1073/pnas.2019284118
